# Impostor Phenomenon in the Nutrition and Dietetics Profession: An Online Cross-Sectional Survey

**DOI:** 10.1101/2021.12.05.21267322

**Authors:** Matthew J. Landry, Dylan A. Bailey, MinJi Lee, Samuel Van Gundy, Audrey Ervin

**Author notes:** **Funding/Financial Disclosures:** No funding or financial disclosures to report. **Availability of Data and Material:** Data and material are available on request from the authors. The material and data that support the findings of this study are available from the corresponding author, MJL, upon reasonable request.

## Abstract

**Background:** Impostor phenomenon (IP) (also known as impostor syndrome) describes high-achieving individuals who, despite their objective successes, fail to internalize their accomplishments and have persistent self-doubt and fear of being exposed as a fraud or impostor. Despite robust literature describing the effects of IP in other health care professions, there is an absence of research within the nutrition and dietetics profession.

**Objective:** To assess the prevalence and predictors of IP within the nutrition and dietetics students and practitioners.

**Design:** An online cross-sectional survey was conducted.

**Participants/setting:** 1,015 students, dietetic interns, and currently practicing and retired nutrition and dietetic technicians registered, and registered dietitian nutritionists provided complete responses.

**Main outcome measures:** Impostor phenomenon was assessed with the Clance Impostor Phenomenon Scale (CIPS). Self-reported Job satisfaction and well-being were assessed using validated scales.

**Statistical analyses:** Descriptive statistics were summarized and reported using frequency counts and percentages. Unadjusted logistic regression models were used to assess the relationship between IP and sociodemographic outcomes, job satisfaction, and well-being.

**Results:** Respondents were primarily female, non-Hispanic White, and practicing dietitians. The average CIPS score was 66.0 ± 16.3 (range 22-99). 64% of survey respondents (n=655) experience intense or frequent IP and 62% (n=628) had a CIPS score ≥62. Older age, educational attainment, professional level, and membership in Academy groups were associated with lower IP scores. Greater Social media use was associated with higher IP scores. Job satisfaction and overall well-being were inversely correlated with impostor phenomenon (p<0.001).

**Conclusions:** Findings from an online survey suggest that a majority of nutrition and dietetics students and practitioners experience IP. Our results reinforce the need to recognize and address this issue by raising awareness, using early prevention methods, and supporting individuals who are younger and/or new to the profession.

## Introduction

Impostor Phenomenon (IP) was first described over 40 years ago by psychology researchers Pauline Clance and Suzanne Imes as persistent cognitions of intellectual phoniness.^1-3^ Simply put, it is the notion that otherwise competent and qualified individuals feel that they have secured esteemed roles or professional positions because of an “oversight” or “plain luck.”^4^ Therefore, they feel like frauds or “impostors.” Impostor phenomenon (IP) is linked with experiencing psychological distress (e.g., depression, anxiety, stress or burnout), increased self-doubt, persistent feelings of failure, strained relationships, as well as significant detrimental outcomes for career advancement and workforce retention.^5, 6^

A recent systemic review of 61 studies and over 14,000 participants found that IP varied widely depending on the screening tool and cutoff used to assess symptoms. Feelings of fraudulence were common across gender, race, age, and a range of occupations though it may be more prevalent and disproportionally impact the experiences of underrepresented or disadvantaged groups.^6^ First-generation college students, academics, marketing managers, chief executive officers, celebrities, and many others report experiencing IP at some time in their life.^3, 6-9^ Psychological characteristics that are associated with increased susceptibility to IP include introversion, trait anxiety, perfectionism, dependence on others for feelings of validation or success, high propensity for shame, high family conflict, generalized anxiety, depressive symptoms, excessive worry, and low self-confidence.^5^

Impostor phenomenon has been described extensively across a wide range of allied health and medicine professionals, including but not limited to nurses and nursing students, physicians, medical students, physician assistants, pharmacy residents, and dental students.^10-16^ Estimates of the prevalence of IP within various healthcare professionals (HCPs) is shown in **Table 1**. HCPs who experience IP report negative psychological outcomes such as increased levels of stress, burnout, as well as decreased job performance and satisfaction over time.^6^ HCPs experiencing IP may overwork and overproduce, which can be counterproductive and lead to burnout. Generally speaking, individuals who experience IP have been found to report less career planning and motivation to lead.^3, 17^ As a result, it is possible HCPs affected by IP may purposefully miss or turn down career advancement opportunities, which could lead to poor career retention.

**Table 1.**
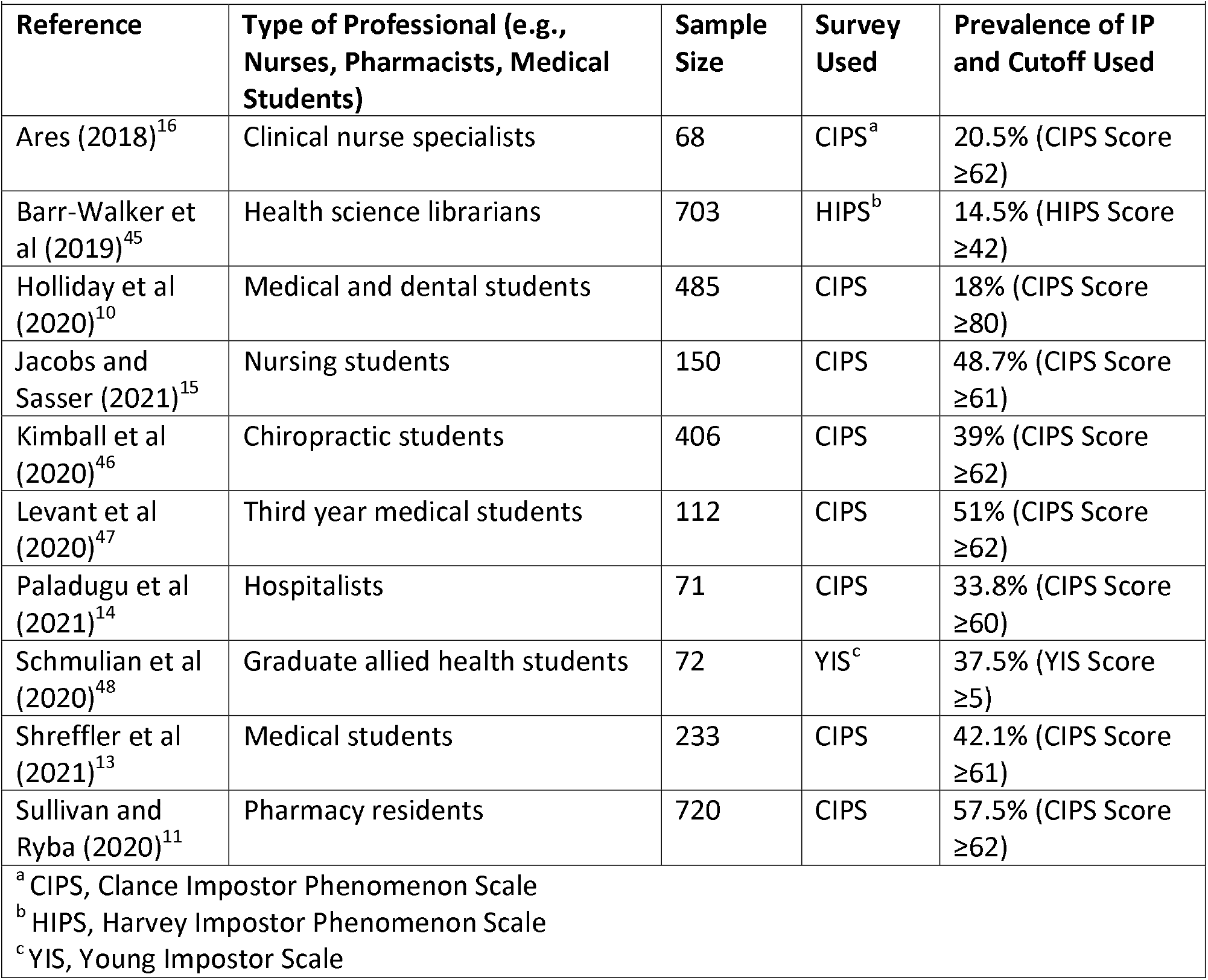
Examples of Prevalence of Impostor Phenomenon within Various Health-Related Professions.

|  |  |  |  |  |
| --- | --- | --- | --- | --- |
| **Reference** | **Type of Professional (e.g., Nurses, Pharmacists, Medical Students)** | **Sample Size** | **Survey Used** | **Prevalence of IP and Cutoff Used** |
| Ares (2018)<sup>16</sup> | Clinical nurse specialists | 68 | CIPS<sup>a</sup> | 20.5% (CIPS Score ≥62) |
| Barr-Walker et al (2019)<sup>45</sup> | Health science librarians | 703 | HIPS<sup>b</sup> | 14.5% (HIPS Score ≥42) |
| Holliday et al (2020)<sup>10</sup> | Medical and dental students | 485 | CIPS | 18% (CIPS Score ≥80) |
| Jacobs and Sasser (2021)<sup>15</sup> | Nursing students | 150 | CIPS | 48.7% (CIPS Score ≥61) |
| Kimball et al (2020)<sup>46</sup> | Chiropractic students | 406 | CIPS | 39% (CIPS Score ≥62) |
| Levant et al (2020)<sup>47</sup> | Third year medical students | 112 | CIPS | 51% (CIPS Score ≥62) |
| Paladugu et al (2021)<sup>14</sup> | Hospitalists | 71 | CIPS | 33.8% (CIPS Score ≥60) |
| Schmulian et al (2020)<sup>48</sup> | Graduate allied health students | 72 | YIS<sup>c</sup> | 37.5% (YIS Score ≥5) |
| Shreffler et al (2021)<sup>13</sup> | Medical students | 233 | CIPS | 42.1% (CIPS Score ≥61) |
| Sullivan and Ryba (2020)<sup>11</sup> | Pharmacy residents | 720 | CIPS | 57.5% (CIPS Score ≥62) |
| <sup>a</sup> CIPS, Clance Impostor Phenomenon Scale<br><sup>b</sup> HIPS, Harvey Impostor Phenomenon Scale<br><sup>c</sup> YIS, Young Impostor Scale |  |  |  |  |

Taken together, there is an accumulating body of work describing the link between impostor feelings and poor job performance, substandard job satisfaction, and burnout among HCPs. Dietetic students and practicing RDNs may display similar types and levels of personality traits, and other psychological characteristics that are associated with increased susceptibility for IP, to the collective body of HCPs reporting IP experiences.^18, 19^ However, studies have not yet evaluated if IP is occurring within dietetic students (e.g., undergraduate or intern), registered dietitian nutritionists (RDN) or nutrition and dietetics technicians, registered (NDTRs).^20^ Given the potential implications on educational and workforce advancement, retention, satisfaction and performance, it critical to understand the level at which IP is occurring in the nutrition and dietetics profession, as well as any factors linked with increased IP susceptibility. Therefore, the primary aim of this study was to describe the prevalence and predictors of IP in the nutrition and dietetics profession.

## Materials and Methods

This cross-sectional, anonymous online survey was administered to nutrition and dietetics practitioners and students and aimed to measure the prevalence of IP and sociodemographic predictors of IP. Procedures were followed in accordance with the ethical standards from the Helsinki Declaration and were approved by the Delaware Valley University Institutional Review Board (protocol #21010, 3 May 2021). All respondents reviewed an informed consent document and agreed to participate.

### Survey Development and Design

The survey consisted of 56 possible questions including demographic questions, the Clance Impostor Phenomenon Scale (CIPS), job satisfaction, burnout and well-being, and social media use. The survey utilized conditional branching to skip blocks of questions that were not pertinent to respondents based on prior responses or indicated professional level (e.g., students, dietetic interns, and retired professionals were not asked questions about job satisfaction). Respondents did not have the option to skip questions that were displayed to them; however, questions that were sensitive in nature had a “prefer not to say” option for respondents to select. It was estimated that respondents would need 15-20 minutes to complete the entire survey.

### Demographics

Data were collected on participant’s age, gender identity, sexual orientation, race, ethnicity, disability status, educational attainment, primary area of practice (for RDNs, NDTRs, and retired)^21^, and membership in the Academy of Nutrition and Dietetics Dietetic Practice Groups (DPGs) and Member Interest Groups (MIGs).^22, 23^

### Impostor Phenomenon

Data on the impostor phenomenon were assessed with the Clance Impostor Phenomenon Scale (CIPS).^2^ Although a gold standard measure is yet to be established, the CIPS is the most cited and utilized measure by practitioners and IP researchers.^24^ The tool is a 20-item survey using a 5-point Likert scale for each item with 1, not at all true; 2, rarely; 3, sometimes; 4, often; and 5, very true. Scores range from 20 to 100, with scores less than 40 indicating few impostor characteristics, scores between 41 and 60 indicating moderate impostor traits, and scores between 61 and 80 representing frequent impostor feelings, and scores of 81 and higher indicating that the respondent possessed intense impostor behaviors. The higher the score, the more frequently and seriously the IP interferes in a person’s life. A CIPS score of ≥62 was used as a threshold to determine respondents demonstrating IP and has been shown to minimize false-positives and false-negatives.^25^ The CIPS has been demonstrated to be reliable with a Cronbach’s of 0.96.^26^ We obtained permission to use the CIPS for this study from the author, Dr. Pauline Rose Clance, via personal communication on January 21, 2021.

### Job Satisfaction

Job satisfaction was assessed using the short-form Minnesota Satisfaction Questionnaire (MSQ).^27^ The MSQ is designed to measure an employee’s satisfaction with his or her current job and consists of three scales: intrinsic satisfaction, extrinsic satisfaction, and general satisfaction. The short-form version of the questionnaire is 20-items which use a 5-point Likert scale for each item with 1, very dissatisfied; 2, dissatisfied; 3, neither; 4, satisfied; and 5, very satisfied. Response choices for all items were summed, yielding a range from 20-100. The higher the total score, the more satisfied an individual is with their job. Only survey respondents self-identifying as a practicing RDN or NDTR completed the MSQ. Using conditional branching, the MSQ was not shown for respondents identifying as a student, intern, or retired RDN or NDTR.

### Burnout and Well-Being

To identify distress in a variety of dimensions (burnout, fatigue, low mental/physical quality of life, depression, anxiety/stress) among all participants the 9-item expanded Well-Being Index (eWBI) was used.^28^ The first seven items are answered using a simple yes/no format. One point is assigned for each “yes.” The expanded index includes one question asking participants to assess their satisfaction with work/internship/school-work life balance and the other on the degree of meaning they derived from work/internship/school-work. Satisfaction with work-life balance was assessed using a 5-point Likert scale (strongly agree; agree; neutral; disagree; strongly disagree). Responses of strongly agree or agree were assigned 1 point, responses of neutral were assigned 0 points, and responses of disagree or strongly disagree are given negative 1 point. Individuals who indicated a low level of meaning in work (response option of a 1 or 2 on a 7-point Likert scale) had 1 point added to their score while those who answered favorably (response option of a 6 or 7 on a 7-point Likert scale) had 1 point subtracted from their score. Those who indicated neutral level of meaning in work (response option of 3 to 5 on the 7-point Likert scale) received 0 points. Response choices for all items were summed, yielding a total score for the eWBI ranging from -2 to 9. Higher scores on the eWBI are indicative of greater risk for distress (i.e., poor well-being).

### Social Media Questions

Seven questions were used to assess social media use. Respondents were asked if they used social media, the number of platforms used, most utilized social media platform, daily amount of time spent on all social media platforms, whether they promoted themselves as a dietitian or dietetics student on their social media account(s), if they compared themselves to other dietitians or dietetics students on social media, and if they felt intellectual phoniness or a persistent feeling of being a fraud compared to other dietitians, nutrition professionals, or influencers on social media.

### Survey Administration

This study utilized a nonrandom, convenience sampling approach. The survey was accessible for two months (May 1 – June 30, 2021). Information about the study was shared through social media groups (e.g., Facebook, Instagram, LinkedIn, etc.) targeting dietetics students, interns, and practicing RDNs/NDTRs and shared via communication channels of Academy Dietetic Practice Groups (DPGs) and Member Interest Groups (MIGs). Directors and administrators of didactic program in dietetics, coordinated program in dietetics, and dietetic internships were asked to pass along information to students and interns within their programs. Lastly, respondents who completed they survey were encouraged to pass along information about the survey to colleagues (snowball recruitment). Eligible respondents who consented to participate, were dietetic students, dietetic interns, practicing RDNs and NDTRs, and retired RDNs and NDTRs.

### Data Management and Statistical Analysis

Survey data were collected and managed using Qualtrics (Qualtrics, Provo, UT, USA). Data from incomplete surveys were not used (N=145, approximately 1% of all collected surveys). Respondents did not have the option to skip questions; therefore, there was no missing data from completed surveys. Categorical variables were described as n (%) and continuous variables were described as mean ± standard deviation. Unadjusted logistic regression models were used to assess the relationship between IP and sociodemographic outcomes. Unadjusted linear regression models were used to examine the relationship between IP and continuous outcomes (job satisfaction from the MSQ and well-being from the eWBI). Data were analyzed in RStudio (Version 1.2.5042, R Core Team, 2019).

## Results

There were 1,015 eligible responses to the survey. Survey respondents were majority RDNs (65%). Most respondents reported being between 25-34 years of age. The sample was primarily female, non-Hispanic, White, and heterosexual (**Table 2**). Nine participants (<1%) reported a chronic disability condition (blindness, deafness, or a severe vision or hearing impairment) and 26 participants (3%) reported a disability condition that substantially limits one or more basic physical activities such as walking, climbing stairs, reaching, lifting, or carrying. Approximately 10% of respondents were a member of at least one Academy DPG and 10% were members of at least one Academy MIG.

**Table 2.** Demographics of Cohort of 1,015 Dietetics Students, Dietetic Interns, and Currently Practicing and Retired Nutrition and Dietetic Technicians Registered and Registered Dietitian Nutritionists Participating in an Online Survey on Impostor Phenomenon.

| Characteristics | n (%) |
| --- | --- |
| Age (years) |  |
| 18-24 | 195 (19.2) |
| 25-34 | 440 (43.3) |
| 35-44 | 194 (19.1) |
| 45-54 | 97 (9.6) |
| 55-64 | 64 (6.3) |
| 65-74 | 24 (2.4) |
| 75+ | 1 (0.1) |
| Gender |  |
| Male | 34 (3.3) |
| Female | 972 (95.8) |
| Non-binary/Nonconforming/Expansive | 5 (0.5) |
| Prefer not to say | 4 (0.4) |
| Ethnicity |  |
| Hispanic/Latino | 87 (8.6) |
| Non-Hispanic/Latino | 928 (91.4) |
| Race |  |
| American Indian or Alaska Native | 8 (0.8) |
| Asian | 48 (4.7) |
| Black or African American | 39 (3.8) |
| Native Hawaiian or Other Pacific Islander | 2 (0.2) |
| White or Caucasian | 853 (84.0) |
| Bi/Multiracial | 65 (6.4) |
| Sexual Orientation |  |
| Heterosexual or straight | 905 (89.1) |
| Gay or lesbian | 17 (1.7) |
| Bisexual | 68 (6.7) |
| Different identity | 12 (1.2) |
| Prefer not to say | 13 (1.3) |
| Educational Attainment |  |
| Some College | 32 (3.2) |
| Associates | 27 (2.7) |
| Bachelors | 453 (44.6) |
| Masters | 442 (43.5) |
| Doctorate | 61 (6.0) |
| Professional Level |  |
| Student (Undergraduate or Graduate) | 170 (16.7) |
| Dietetic Intern | 151 (14.9) |
| Practicing NDTR <sup>a</sup> | 18 (1.8) |
| Practicing RDN <sup>b</sup> | 658 (64.8) |
| Retired NDTR or RDN | 18 (1.8) |
| Chronic Disability Condition <sup>c</sup> | 9 (0.9) |
| Disability Substantially Limiting Basic Physical Activities <sup>d</sup> | 26 (2.6) |
| Member of an Academy <sup>e</sup> Dietetic Practice Group | 105 (10.3) |
| Member of an Academy <sup>e</sup> Member Interest Group | 105 (10.3) |
| <sup>a</sup> NDTR, Nutrition and Dietetics Technician Registered<br><sup>b</sup> RDN, Registered Dietitian Nutritionist<br><sup>c</sup> Blindness, deafness, or a severe vision or hearing impairment<br><sup>d</sup> Disability condition that substantially limits one or more basic physical activities such as walking, climbing stairs, reaching, lifting, or carrying.<br><sup>e</sup> Academy of Nutrition and Dietetics |  |

Of practicing RDNs and NDTRs, the majority (45%) have been practicing for less than five years, 19% have 5-9 years of practice, 20% have 10-19 years, and 16% have been practicing 20 or greater years. Of retired RDNs and NDTRs, the majority (89%) had practiced 20 or greater years prior to retirement. The majority (54%) of practicing RDNs and NDTRs reported practicing in clinical nutrition with 25%, 19% and 10% in acute care/inpatient care, ambulatory care, and long-term care, respectively. The remaining practicing RDNs and NDTRs reported practice areas in community nutrition (18%), education and research (14%), consultation and business (10%), and food and nutrition management (5%). Within retired RDNs and NDTRs, 56% reported working in clinical nutrition prior to retirement with 28%, 17% and 11% in acute care/inpatient care, ambulatory care, and long-term care, respectively. The remaining retired RDNs and NDTRs reported practice areas in community nutrition, 11% in education and research, and 6% in consultation and business prior to their retirement.

### Prevalence of IP

Across all nutrition and dietetics professional levels, the average Clance Impostor Phenomenon score (CIPS) was 66.0 ± 16.3 (range 22-99) (**Table 3**). Intense IP experiences were reported by 21% of respondents and 43%, 28%, and 8% of respondents reported frequent, moderate, and few IP experiences, respectively. Using the CIPS threshold score of 62 out of 100, 628 respondents (62%) were identified as having IP. Prevalence of IP experiences stratified by professional level are shown in **Table 3** and displayed in **Figure 1**.

**Table 3.** Prevalence of Impostor Phenomenon Experiences in Nutrition and Dietetics by Professional Level.

|  |  |  |  |  |  |  |
| --- | --- | --- | --- | --- | --- | --- |
| **Impostor Phenomenon Experiences** | **Student (n=170)** | **Dietetic Intern (n=151)** | **NDTR <sup>a</sup> (n=18)** | **RDN <sup>b</sup> (n=658)** | **Retired NDTR/RDN (n=18)** | **Total (n=1,015)** |
| **Few** | 7 (4.1%) | 7 (4.6%) | 0 (0.0%) | 58 (8.8%) | 6 (33.3%) | 78 (7.7%) |
| **Moderate** | 31 (18.2%) | 36 (23.8%) | 4 (22.2%) | 206 (31.3%) | 5 (27.8%) | 282 (27.8%) |
| **Frequent** | 80 (47.1%) | 69 (45.7%) | 11 (61.1%) | 276 (41.9%) | 4 (22.2%) | 440 (43.3%) |
| **Intense** | 52 (30.6%) | 39 (25.8%) | 3 (16.7%) | 118 (17.9%) | 3 (16.7%) | 215 (21.2%) |
| <sup>a</sup> NDTR, Nutrition and Dietetics Technician Registered |  |  |  |  |  |  |
| <sup>b</sup> RDN, Registered Dietitian Nutritionist |  |  |  |  |  |  |

**Table 4.**
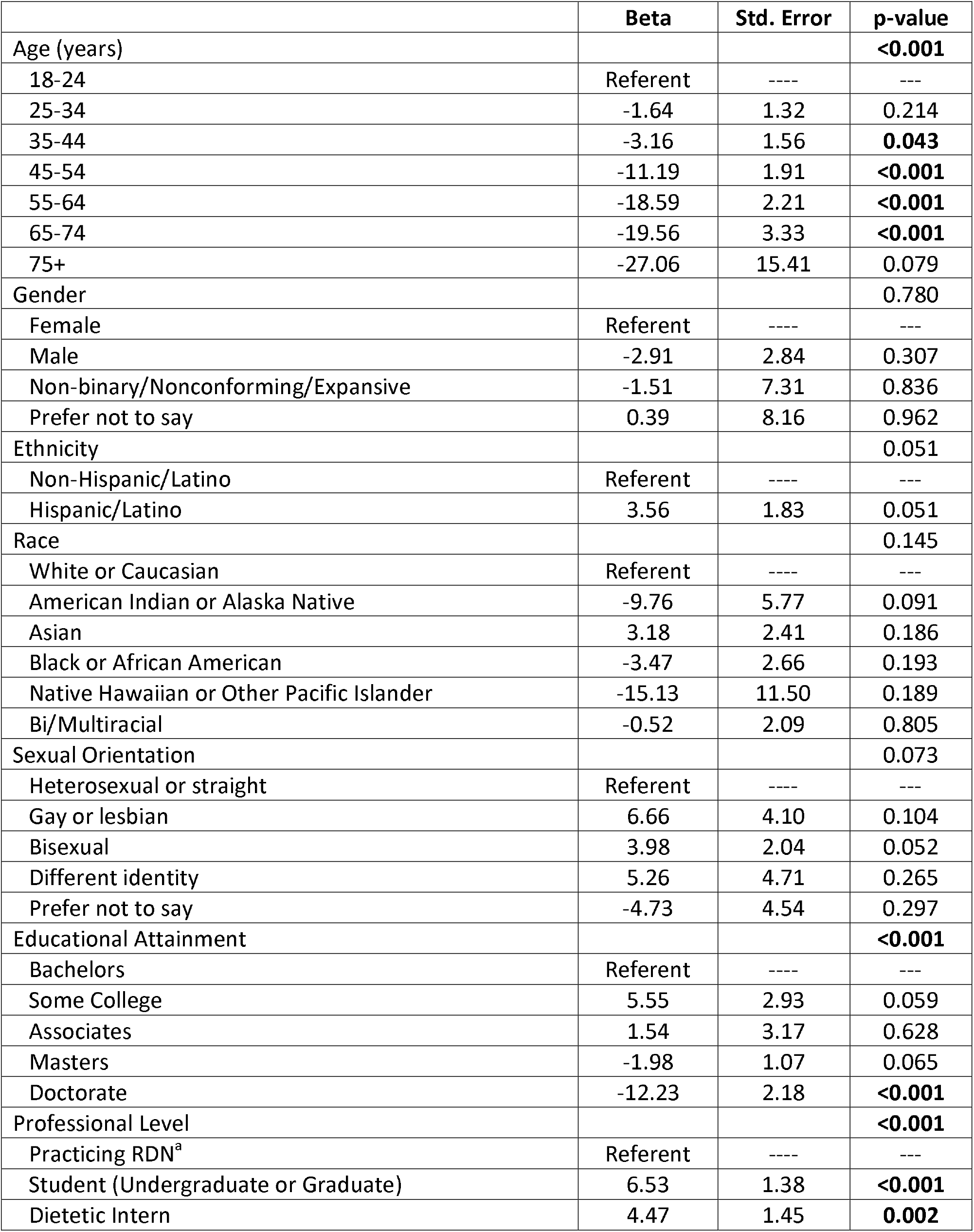
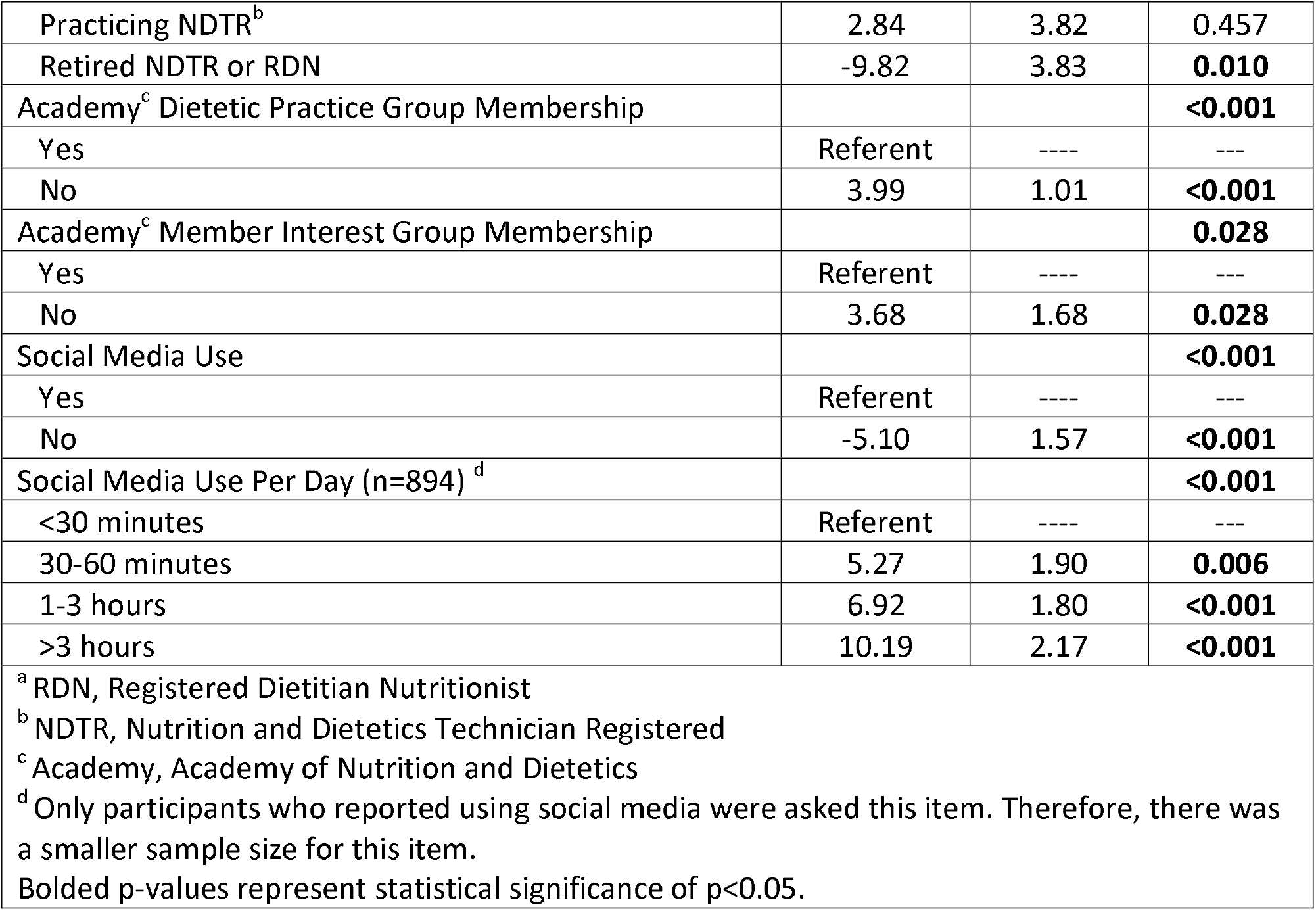
Demographic Predictors of Impostor Phenomenon in the Nutrition and Dietetics Profession (n=1,015)

**Figure 1.**
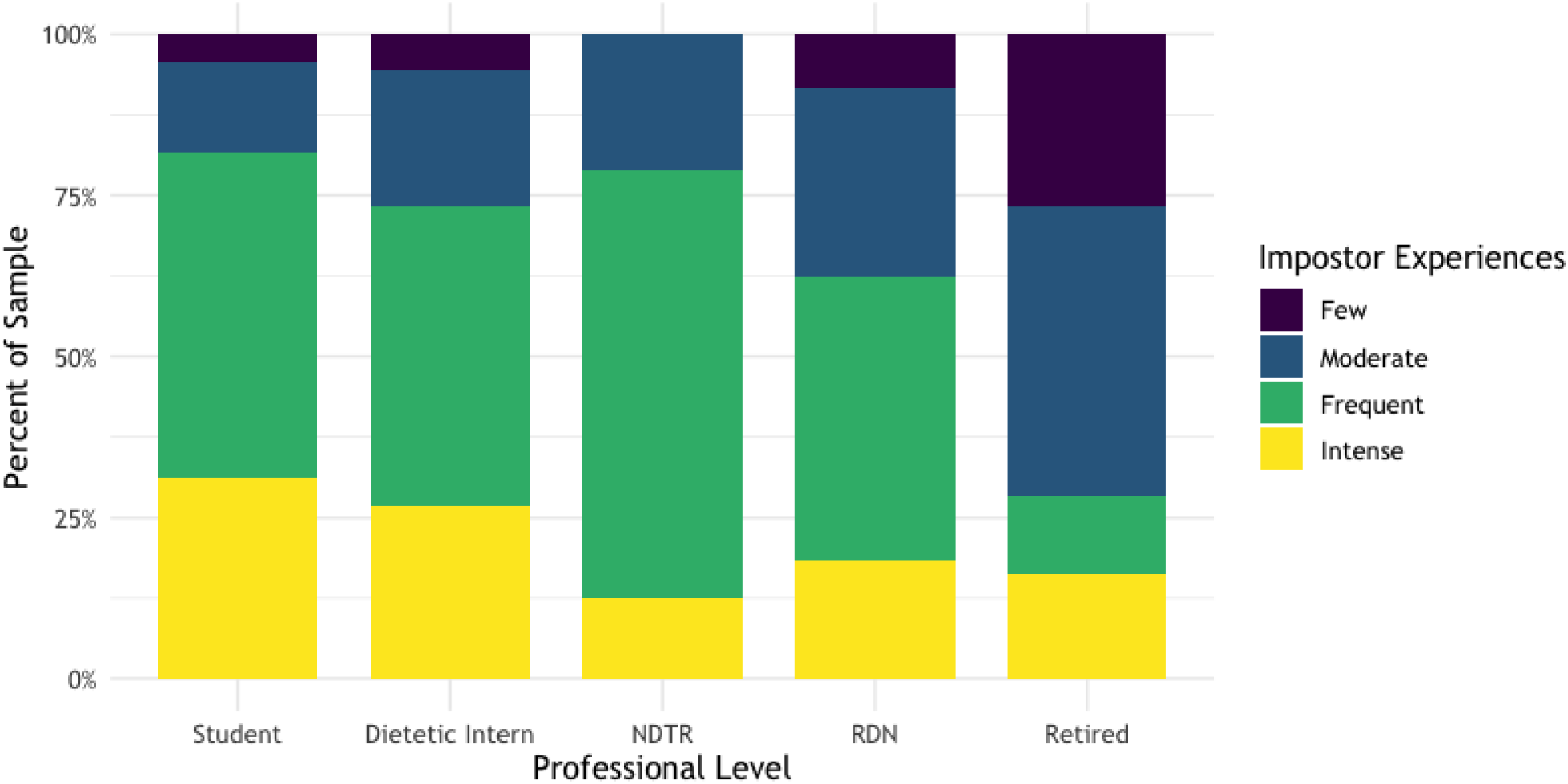
Impostor Phenomenon by Professional Level (n=1,015). Data on the impostor phenomenon were assessed with the Clance Impostor Phenomenon Scale. Abbreviations: NDTR, Nutrition and Dietetics Technician Registered; RDN, Registered Dietitian Nutritionist

### Demographic Predictors of Impostor Phenomenon

There were several demographic predictors that were associated with higher CIPS scores (i.e., more likely to experience impostor experiences). Age was a significant predictor of CIPS scores (p<0.001). Significant differences were found in individuals 35-64 years of age compared to 18-24 years of age respondents (p<0.001). There was a protective effect of age on CIPS scores, as older respondents had lower CIPS scores. Educational attainment was a significant predictor of CIPS scores; however, there was only a between-group difference of those with a bachelor’s degree and those with a Doctorate. Professional level was a significant predictor of CIPS scores. Students (undergraduate or graduate) had 6.5 points lower CIPS scores (p<0.001) compared to practicing RDNs and interns had 4.4 points lower CIPS scores (p=0.002) compared to practicing RDNs. However, retired NDTRs and RDNs had 10 points lower CIPS scores compared to practicing RDNs. There was no significant difference in CIPS scores between practicing RDNs and practicing NDTRs. Membership in an Academy DPG or MIG was found to be associated with lower CIPS scores (p<0.001 and p<0.028, respectively). Compared to members, respondents who were not a member of any DPG had 4 points higher CIPS scores. A similar association was found for respondents who were not part of a MIG. Use of social media was significantly associated with higher CIPS scores (p<0.001). There was also stepwise positive relationship between amount of social media use per day and higher CIPS scores (p<0.001). Participants who reported the highest social media use (>3 hours per day) had 10 points higher CIPS scores compared to those who used social media for <30 minutes per day. Gender, ethnicity, race, and sexual orientation were not significant predictors of impostor phenomenon within the study.

### Well-Being

Average expanded Well-Being Index (eWBI) score was 3.3 ± 2.4 (range -2, 9). Among all participants, there was an inverse relationship observed with IP and eWBI score. For every one-unit increase in IP, risk of poor well-being went up by 0.07 (p<0.001).

### Job Satisfaction

Within practicing RDNs and NDTRs, average job satisfaction on the short-form Minnesota Satisfaction Questionnaire was 73.3 ± 11.7 (range 33-100). For everyone one-unit increase in IP, job satisfaction was associated with a decrease of 0.16 (p<0.001).

### Social Media Use

The majority (88%) of survey respondents reported currently using social media (**Table 5**). Of those using social media, more than 61% spend more than one hour on social media per day. Most respondents reported having two to three active social media accounts with Facebook and Instagram as the social media channels respondents were most active on. Less than one-third of respondents use social media as a tool to promote themselves as a dietitian or dietetics student. However, nearly 40% of respondents compare themselves to other dietitians or dietetics students on social media at least half the time, and 44% reported feeling intellectual phoniness or a persistent feeling of being a fraud compared to other dietitians, nutrition professionals, or influencers on social media.

**Table 5.** Social Media Use Among Dietetics Students, Dietetic Interns, and Currently Practicing and Retired Nutrition and Dietetic Technicians Registered and Registered Dietitian Nutritionistsa.

|  | % of sample |
| --- | --- |
| **Do you currently use or are you active on social media platform(s)? (n=1,015)** |  |
| Yes | 88.1% |
| No | 11.9% |
| **How many social media sites do you have accounts with?** |  |
| 1 | 3.8% |
| 2-3 | 45.6% |
| 4-5 | 33.8% |
| 6 or more | 16.8% |
| **How much time do you collectively spend on social media per day?** |  |
| <30 minutes | 10.3% |
| 30-60 minutes | 28.6% |
| 1-3 hours | 47.7% |
| >3 hours | 13.4% |
| **Which social media channel are you most active on** |  |
| Facebook | 37.6% |
| Twitter | 2.6% |
| Instagram | 45.4% |
| LinkedIn | 2.5% |
| Pinterest | 0.7% |
| YouTube | 5.4% |
| TikTok | 5.9% |
| **Do you use social media to promote yourself as a dietitian or dietetics student?** |  |
| Yes | 30.9% |
| No | 69.1% |
| **Do you compare yourself to other dietitians or students on social media?** |  |
| Always | 15.8% |
| Most of the time | 12.1% |
| About half the time | 11.2% |
| Sometimes | 41.8% |
| Never | 19.1% |
| **Do you feel intellectual phoniness or a persistent feeling of being a fraud compared to other dietitians, nutrition professionals, or influencers on social media?** |  |
| Always | 13.7% |
| Most of the time | 16.5% |
| About half the time | 14.2% |
| Sometimes | 29.0% |
| Never | 26.7% |
| <sup>a</sup> n=894 unless otherwise denoted |  |

## Discussion

In this online cross-sectional survey of 1,015 nutrition and dietetics professionals, 62% were identified as having IP (CIPS score ≥62) and 64% reported experiencing intense or frequent IP. Job satisfaction and overall well-being scores were also found to be inversely associated with IP scores. Older age, educational attainment, professional level, and membership in Academy groups were associated with lower IP scores. Greater Social media use was associated with higher IP scores. Compared to other healthcare professionals with documented IP experiences (Table 1), this study is one of the largest to suggest that nutrition and dietetics professionals present with the highest prevalence of IP across other HCP domains and professional levels.

In this study, impostor experiences declined with increasing age; however, nutrition and dietetics professionals at all stages of the education pipeline and career ladder reported experiencing impostorism. A protective age effect is not always present within the literature.^6^ In this regard, a practitioner’s time and familiarity in their current position may be a stronger predictor of fewer impostorism experiences. This is partially corroborated by our findings that greater education, experience in the field, and professional level (i.e., student vs intern vs RDN) have a protective effect on IP symptomology. As dietetic educational programs are the gatekeepers to the profession, it is important to acknowledge that they may simultaneously foster insecurities and limit efforts to recognize and support those struggling with them. Additionally, training educators on the early identification of distress due to IP symptomology and deployment of psychologically appropriate coping strategies, encouragement of more realistic perceptions via modelling, and normalizing performance concerns from students may be warranted.

Programs and internships can be adapted to incorporate strategies and mentoring networks to allow for students and interns to overcome persistent feelings of self-doubt or overwhelming drive for perfectionisim.^20, 29^ For example, students may benefit from openly discussing imposter feelings, context-specific fears or concerns, and perfectionism during orientation and/or peer discussions. For practitioners, professional development programs and mentoring networks can be utilized to mitigate attributing success to external factors or setbacks as evidence of professional inadequacy or lack of competence. Peer-to-peer workshops have been utilized by other health professionals as a means to discuss discipline-specific strategies on how to address IP at the individual, peer, and professional institution levels and could be implemented within the nutrition and dietetics profession.^30, 31^

Gender was not found to not be predictive of greater IP experiences. A systematic review of 33 articles found mixed results when comparing prevalence of impostor syndrome by gender.^6^ Half of studies found that women reported greater impostor feelings while the other half of studies found that there were no differences in prevalence. Despite the inconclusive evidence of gender effects, both genders experience impostorism. Gender identity is an important consideration when developing strategies to overcome impostor feelings as previous research has found that men and women cope differently with these experiences.^32, 33^ Sexual orientation was also not predictive of greater IP experiences. Interpretation of gender and sexual orientation as non-significant predictors of IP experiences within this sample should be made with caution. Respondents were primarily cis-gender, heterosexual females, and the sample included few respondents who self-identified as part of a gender- or sexual-minority group.

Race and ethnicity were not predictive of greater IP experiences in this study. However, within the literature racial and ethnic minority groups have been found to have higher rates of IP.^6, 34-36^ It should also be noted that the racial and ethnic categorizations used within the study may overlook differences among heterogenous diverse racial and ethnic groups. Mullangi and Jagsi (2019) have suggested that IP is but a symptom where inequity is the true underlying disease.^37^ The root cause of higher rates within underrepresented groups is likely a result of inequities that exist within the structural and social environments. Acknowledging that the dietetics profession is predominantly able-bodied, cisgender, heterosexual, White, and female, improvements in diversity across the dietetics profession is an important first step, as it provides diverse role models needed to encourage students to enter and remain within the profession.^38, 39^ Considering the important role of the environment in provoking IP feelings among nutrition and dietetics professionals may offer more structural and effective solutions, but requires additional research.^4^

Membership in Academy DPGs and MIGs were found to be protective against IP experiences. Academy DPGs enable students and practitioners to engage with like-minded colleagues around a specialized area of practice.^22^ Through active participation within the DPG, it may enable members to seek specific supportive guidance and improve their comfortability and competence with an area of practice. Similarly, Academy MIGs which reflect characteristics of the Academy’s membership and the public it serves (i.e., Latinos and Hispanics in Dietetics and Nutrition (LAHIDAN) and Cultures of Gender and Age (COGA)) may play a critical role in providing mentorship and networking opportunities for students and practitioners when local networkers lack diversity.^23^

Respondents who reported greater IP experiences were associated with having poorer well-being and job satisfaction. Nutrition and dietetics professionals who experience IP experiences may not develop their professional potential. Those surveyed had an average well-being score of 3.3 and scores on the eWBI that are ≥2 have been associated with greater risk for adverse outcomes including burnout, severe fatigue, suicidal ideation, and poor overall quality of life among employed US adults.^28^ Job satisfaction ranged in the sample, however, the average was only 73 out of a possible 100. Impaired job performance, job satisfaction, and burnout have all been associated with IP based on findings from a systematic review.^6^ It is unclear whether experiences of impostorism impact practice or the outcomes for clients and patients. It is important to note that the current survey was collected in Spring 2021, a year after the onset of the COVID-19 pandemic. Healthcare providers have felt greater stress and burnout, and increased job dissatisfaction and workload during the pandemic.^40, 41^ The COVID-19 pandemic caused many changes to dietitian job responsibilities and modalities of care.^42^ This may have impacted how survey respondents answered self-reported measures of well-being and job satisfaction than they would have prior to the onset of the pandemic. These findings suggest that greater organizational support be provided to employees by managers and administrators to understand why self-doubt develops, how it is sustained, and how nutrition and dietetics practitioners can develop resilient management strategies to mitigate the negative effects of IP.

Social media has the potential to be a positive influence on nutrition if reputable sources are successfully used to spread accurate, evidence-based nutrition information, find healthy recipes, and create safe online environments for sharing.^43^ The ready availability of social networking sites also provide abundant opportunities for social comparisons.^44^ In this study, a majority of those surveyed reported being active on social, in some cases, spending several hours per day on social media sites. During this time, many respondents said that they compare themselves to other RDNs on social media. Further research is needed to elucidate RDN behaviors on social media and how this contributes to their self-esteem and feelings of impostorism.

### Strengths & Limitations

This study did not aim to test a hypothesis about a population, but rather develop an initial understanding of the prevalence of IP within the nutrition and dietetics profession. The survey relied on self-reported measures of IP, well-being, job satisfaction, and social media usage. Social desirability bias may have impacted responses. In addition, there is the potential for self-selection bias, in that individuals within the nutrition and dietetics profession with a particular interest in IP may have been more likely to respond, and for nonresponse bias. Thus, results should be interpreted accounting for these potential biases.

Because the survey was widely distributed, it is difficult to determine the response rate of this survey and, thus, generalizability to the greater nutrition and dietetics population. However, compared to prior studies of IP within health-related professionals this study is among the largest to report IP prevalence. Additionally, prior studies in other professions largely survey only students. This study examined individuals across the entire professional continuum which allowed for a greater understanding of prevalence of IP at various career stages. This study provides an initial benchmark for which a more robust and widely circulated survey could be conducted allowing for a more generalizable assessment within the profession.

## Conclusion

In a non-representative online survey of nutrition and dietetics students and professionals, greater than 60% of respondents reported experiencing IP. Results of this study reinforce the need for recognizing and addressing this issue by raising awareness, normalizing IP discussions, using early prevention methods, and supporting individuals who are younger and/or new to the profession.

Additional research on preventative strategies and early interventions is needed to understand if it is possible to prevent the initial symptoms of IP across the professional level. Identifying the appropriate tools for promoting self-awareness, self-confidence and critical reflection of work and/or education-based experiences within the nutrition and dietetics profession is required in addition to exploring factors that influence confidence and resilience.

Impostor phenomenon should be viewed not just as a personal challenge that someone must overcome but a possibly widespread occurrence within nutrition and dietetics practitioners at various stages of their career. An important implication for practice is the need for future research in this area to identify if and how impostorism affects practice and the outcomes for clients/patients.

## Supporting information

Reporting Checklist

## Data Availability

All data produced in the present study are available upon reasonable request to the authors

